# Dietary Practice and its Association with Glycemic Control among Individuals with Type 2 Diabetes Mellitus in Ethiopia: A Multi-Center Cross-Sectional Study

**DOI:** 10.1101/2023.08.26.23294671

**Authors:** Firegenet Asnake Kitaw, Muluken Zeleke Megiso, Indris Ahmed Yesuf, Bersabel Hilawi Tewodros, Yohannes Shiferaw Shaweno, Blen Solomon Teklu, Mefthe Fikru Berhanu, Edengenet Solomon Weldesenbet, Tigist Workneh Leulseged

**Author notes:** Corresponding author: Firegenet Asnake Kitaw.

## Abstract

**Background:** Type 2 Diabetes Mellitus (T2DM) is a chronic metabolic disorder that needs a comprehensive management plan. The integral role of nutrition therapy in diabetes management is getting special attention in guidelines though the practice is in a primitive stage, especially in resource limited settings where lifestyle modifications have a large role in the overburdened healthcare system. Understanding the current dietary practice and its effect on disease control in settings where a tertiary level care is provided is a critical step in providing targeted intervention. Therefore, the aim of the study was to assess dietary practice and its association with level of glycemic control among individuals with T2DM who were on follow-up at two large tertiary hospitals in Ethiopia.

**Methods:** A hospital based cross-sectional study was conducted from January to March 2023 among 314 systematically selected individuals with T2DM who were on follow-up at diabetes clinics of St. Paul’s Hospital Millennium Medical College and Tikur Anbessa Specialized Hospital. Data was collected using a pre-tested structured questionnaire and summarized using frequency and median (interquartile range). To examine the association of dietary practice with level of glycemic control, a binary logistic regression model was run at 5% level of significance where Adjusted Odds Ratio (AOR) and 95% CI for AOR were used to interpret the results.

**Results:** From the 314 participants, 146 (46.5%) patients had adequate knowledge regarding the recommended dietary practices and only 42 (13.4%) of the individuals practiced a healthy diet. A total of 107 (34.1%, 95% CI=29.0%-39.2%) had optimal glycemic control. Poor dietary practice (AOR=7.93, 95% CI=2.63-23.89, p<0.001), obesity (AOR=2.74, 95% CI=1.05-7.18%, p=0.04), and taking combination oral anti-diabetic drugs (AOR=6.22, 95% CI=3.05-12.69, p<0.001) were significantly associated with suboptimal glycemic control.

**Conclusions:** Dietary knowledge and practice among individuals with T2DM were very low, as is the level of glycemic control, which are similar to studies conducted years back, indicating a lack of improvement in the desired behavior over time. Poor dietary practice in turn was associated with suboptimal glycemic control. It is important to target interventions that enhance the understanding and application of dietary practice in these individuals.

## BACKGROUND

Type 2 diabetes mellitus (T2DM) is a chronic metabolic disorder characterized by hyperglycemia caused by the body’s failure to produce or utilize insulin adequately (1). The global prevalence of T2DM has risen in recent years, with an estimated 537 million adults living with the disease by 2021. Africa has the fastest growing prevalence of T2DM, with 24 million people living with the disease in 2021, and the figure is predicted to rise to 55 million by 2045, the largest growth of any International Diabetes Federation (IDF) region. In Ethiopia, 1.9 million people were estimated to live with the disease in the year 2021 (2).

Without early diagnosis and management, T2DM can lead to serious long-term complications including cardiovascular disease, stroke, chronic kidney disease, damage to the nerve, leg ulcer, and increased risk of death (2,3). The combination of rise in incidence, morbidity and mortality rates related to diabetes presents a big challenge to the health-care system and lowers the standard of life of those affected (4). Therefore, achieving optimal glycemic control is vital (1-2,5). However, despite the growing attention given to the management of chronic medical illness, including diabetes, and the numerous interventions designed to help individuals achieve their treatment goals and live a quality life, the vast majority of these people have suboptimal glycemic control. This is true for both developing and developed nations, with multiple studies revealing inadequate glycemic control in up to three-quarters of patients (6-10).

To achieve optimal glycemic control, a comprehensive management plan that includes dietary changes, exercise, and weight loss in addition to standard pharmacological therapy is required (1-2,5). Dietary management is one component of the management that should be followed over long term and is expected to lower hemoglobin A1C levels by 1%-2%, thereby preventing or delaying micro and macro-vascular complications. Despite the existence of published standards for optimum diabetes treatment, evidence suggests that translating nutrition guidelines into daily routine can be challenging for the majority of people with diabetes (5,11). Numerous researches have also found that 50-80% of the studied population had poor dietary practices and also knowledge of the recommended dietary practices (12-19). The lack of adherence to the recommended dietary practice, in turn, is directly associated with an increased risk of suboptimal glycemic control. It has been reported that those with poor dietary habits have a fivefold increased risk of having suboptimal glycemic control, necessitating the escalation of pharmacologic treatment and thus increasing the risk of disease complication and drug side effects on the patient (19-22).

In resource-constrained countries such as Ethiopia, the cost of healthcare is often high, necessitating a significant emphasis on preventive aspects of disease management such as lifestyle modifications, as caring for those with complications can be costly or unavailable, resulting in poor quality of life and an increased risk of mortality. As a result, understanding the current practice and its effect on disease control in settings where a tertiary level care is provided and where dietary practice is expected to be better is crucial to understanding the current disparity in these people’s dietary habits and hence is a critical step in providing targeted intervention. Therefore, the aim of this study was to assess the dietary practice of individuals with T2DM and its association with level of glycemic control among those who were on follow-up at two large tertiary hospitals in Ethiopia.

## METHODS AND MATERIALS

### Study Design and Setting

A hospital based Cross-sectional study was conducted from January to March, 2023 at the two largest tertiary hospitals in Ethiopia; St. Paul’s hospital millennium medical college (SPHMMC) and Tikur Anbessa Specialized Hospital (TASH) which are located in Addis Ababa, the capital city of Ethiopia. Both hospitals have endocrine clinics where patients are followed by internal medicine residents, internists and/or fellow and senior endocrinologists. The majority of patients in these clinics are diabetic, accounting for 80-90% of total patients attending the endocrine outpatient department, and an average of 360 diabetic patients per month at SPHMMC and 120 patients per month at TASH were seen in the three months preceding the study.

### Population and Eligibility

The study included non-pregnant adults with a confirmed diagnosis of T2DM who were on follow-up at the endocrine clinics of the hospitals from January to March 2023, had been on follow-up for at least six months, and were in stable medical condition at the time of the study.

### Sample Size Determination and Sampling Procedure

Sample size for the descriptive objective of the study (to determine the level of glycemic control) was calculated by using single population proportion formula by taking proportion of suboptimal glycemic control as 72%, 5% level of significance, 5% margin of error and the calculated sample size was 310. And sample size for the inferential objective (association of dietary habit with glycemic control) was calculated using double population proportion formula with the following assumptions; 95% confidence interval, power of 80%, the proportion of individuals with T2DM with good dietary habit who had suboptimal glycemic control as 32%, proportion of individuals with T2DM with bad dietary habit who had suboptimal glycemic control as 68%, and a non-response rate of 10% and the calculated sample size was 70 (10). By taking the largest sample size from the two and adjusting for non-response rate of 10%, the final sample size was 341. This was then proportionally allocated to the two hospitals; (257 from SPHMMC and 84 from TASH). To select the study participants, a systematic random sampling method using the k^th^ interval of 2 was employed.

### Operational Definitions

#### Glycemic control

According to American Diabetes Association (ADA) criteria, optimal glycemic control is defined as a three-month average fasting blood sugar of 80-130 mg/dl. Any result out of this range signifies suboptimal glycemic control (23).

#### Dietary practice

Practice was assessed using 12 questions and the total response was classified in to three groups based on the modified Bloom’s cut-off point as good (80-100%), moderate (60-79.9%) and poor (<60%) practice (9,10,14,19,24)

#### Dietary knowledge

Knowledge was assessed using eight questions and the total response was classified in to three groups based on the modified Bloom’s cut-off point as adequate (80-100%), moderate (60-79.9%) and inadequate (<60%) knowledge (9,10,14,19,24).

#### Physical activity

According to ADA recommendation on physical activity for individual with diabetes, it is classified as (23);

- **Optimal (Moderate/ vigorous) activity:** Prolonged, rhythmic activity using large muscle groups (e.g., walking, cycling and swimming), at least 150 min/week, 3-7 days/weeks
- **Sub-optimal:** Any activity less than what is stated above.

### Data Collection and Quality Assurance

A pre-tested structured interviewer administered questionnaire was used to collect data on sociodemographic, clinical characteristics, dietary knowledge and practice, and glycemic control level of the participants. Data was collected by four trained General practitioners who were working at chronic follow-up clinics of the hospitals.

Dietary Knowledge was assessed using eight questions that were adopted from other literature and contextualized into the local context. Each question was answered using the “Yes”, “No” and “I don’t know” options. A correct answer (“Yes”) was assigned 1 point and an incorrect//unknown answer (“No” and “I don’t know”) were assigned 0 points. The total score for each patient on the eight questions was calculated and changed to percentage. After that the score of each participant was changed into a three-category response based on modified Bloom’s cut-off point, as adequate (if score was between 80 and 100%), moderate (if the score was between 60 and 79.9%), and inadequate (if the score was less than 60%) knowledge. Dietary practice was assessed using 12 questions with two response categories (“Yes” and “No”), where those who answered “Yes” were given a score of 1 and those who answered “No” were given a score of 0. The total score for each patient on the 12 items was calculated and changed to percentage. Then, modified Bloom’s cut-off point was used to categorize the response as good (80-100%), moderate (60-79.9%) and poor (<60%) practice.

Weight and height were measured by the data collectors by following the proper methods of measurements. The body mass index (BMI) of each participant was then calculated using the formula: BMI = weight (kg) / height (m^2^) and participants were classified according to the WHO International classification of adult weight. Finally, the three months average FBS was calculated for each patient from the results attached on their chart.

### Statistical Analysis

The characteristics of the participants were presented using frequency with percentages for categorical variables. For numeric variables, median (with interquartile range) was used due to the skewed distribution of the numeric data, as evidenced by a significant result on the Kolmogorov-Smirnov test of normality (p-value <0.0001).

To examine the association of dietary practice and the other factors with glycemic control, a binary logistic regression model was used. At a p-value of < 0.25, a univariate analysis was run to assess the relationship of each factor with level of glycemic control. Variables with significant association were then fitted into the final multivariable model where Adjusted Odds ratio (AOR) and 95% confidence interval for AOR were used to interpret significant results at a p-value of <0.05. The final model’s adequacy was tested using Hosmer and Lemeshow test, and the result showed that the data fitted the model very well (X^2^_(8)_ =6.67 and p-value=0.573). All data management and analysis were done using SPSS software Version 25.0.

## RESULTS

### Socio-demographic Characteristics

Out of the total of 341 participants, 314 completed the interview and were included in the analysis, making a response rate of 92.1%. The median age of the participants was 56.0 years (IQR, 50.0-64) with the majority being older than 40 years, with most 50-59 years old (35.0%). The study included a nearly equal proportion of males (50.3%) and females (49.7%). Only 26 (8.3%) of the participants were unable to read or write; the remainder have formal schooling experience ranging from primary to master’s level. The majority (61.8%) were followers of Orthodox Christian religion. Over a third (71.3%) were from Addis Ababa, while 71 (22.6%) were residents of the Oromia region. **(Table 1)**

**Table 1:** Socio-demographic characteristics of individuals with T2DM on follow up at two tertiary hospitals Ethiopia, 2023 (n=314)

| Variable |  | Frequency | Percentage |
| --- | --- | --- | --- |
| Age category (in years) | 22-39 | 10 | 3.2 |
|  | 40-49 | 64 | 20.4 |
|  | 50-59 | 110 | 35.0 |
|  | 60-69 | 87 | 27.7 |
|  | 70-82 | 43 | 13.7 |
| **Sex** | Male | 158 | 50.3 |
|  | Female | 156 | 49.7 |
| **Marital status** | Single | 8 | 2.5 |
|  | Married | 239 | 76.1 |
|  | Divorced | 22 | 7.0 |
|  | Widowed | 45 | 14.3 |
| **Educational level** | Cannot read and write | 26 | 8.3 |
|  | Primary | 143 | 45.5 |
|  | Secondary | 57 | 18.2 |
|  | Diploma and above | 88 | 28.0 |
| **Religion** | Orthodox | 194 | 61.8 |
|  | Muslim | 73 | 23.2 |
|  | Protestant | 46 | 14.6 |
|  | Catholic | 1 | 0.3 |
| **Place of residence** | Addis Ababa | 224 | 71.3 |
|  | Oromia | 71 | 22.6 |
|  | Others <sup>a</sup> | 21 | 6.7 |

<sup>a</sup> Amhara, SNNP, Dire Dawa, Afar, and Sidama

### Behavioral and Clinical Characteristics

One or more comorbid illnesses were diagnosed in 277 (88.2%) patients, with hypertension accounting for 246 (78.3%) cases, followed by neurologic conditions in 126 (40.1%), renal condition in 69 (22.0%), cardiac disease in 47 (15.0%), and dyslipidemia in 29 (9.2%).

At the time of the study, almost three-quarters were above normal weight; 135 (43.0%) were overweight and 83 (26.4%) were obese. Two hundred forty-six (78.3%) participants claimed to exercise sub-optimally. Twelve (3.8%) were smokers, and 49 (15.6%) chewed Khat.

The median disease duration following T2DM diagnosis was 8.6 years (IQR, 6.0-11.0 years), and nearly half of the patients (49.7%) had diabetes for 6-10 years. At the time of the study, all of the participants were taking oral anti-diabetic medication; 32 (10.2%) were taking Glibenclamide, 80 (25.5%) were taking Metformin, and the remaining 202 (64.3%) were taking both prescriptions. **(Table 2)**

**Table 2:** Behavioral and clinical characteristics of individuals with T2DM on follow up at two tertiary hospitals in Ethiopia, 2023 (n=314)

| **Variable** |  | **Frequency** | **Percentage** |
| --- | --- | --- | --- |
| **Comorbid illness** | No | 37 | 11.8 |
|  | Yes | 277 | 88.2 |
| **Hypertension** | No | 68 | 21.7 |
|  | Yes | 246 | 78.3 |
| **Dyslipidemia** | No | 285 | 90.8 |
|  | Yes | 29 | 9.2 |
| **Cardiac** | No | 267 | 85.0 |
|  | Yes | 47 | 15.0 |
| **Neurology** | No | 188 | 59.9 |
|  | Yes | 126 | 40.1 |
| **Renal** | No | 245 | 78.0 |
|  | Yes | 69 | 22.0 |
| **Asthma** | No | 299 | 95.2 |
|  | Yes | 15 | 4.8 |
| **HI** | No | 301 | 95.9 |
|  | Yes | 13 | 4.1 |
| **BMI Category** | Underweight | 7 | 2.2 |
|  | Healthy weight | 89 | 28.3 |
|  | Overweight | 135 | 43.0 |
|  | Obese | 83 | 26.4 |
| **Exercise** | Moderate/ vigorous | 68 | 21.7 |
|  | Sub-optimal | 246 | 78.3 |
| **Smoking** | No | 302 | 96.2 |
|  | Yes | 12 | 3.8 |
| **Khat chewing** | No | 265 | 84.4 |
|  | Yes | 49 | 15.6 |
| **Disease duration (in years)** | 1-5 | 69 | 22.0 |
|  | 6-10 | 156 | 49.7 |
|  | ≥ 11 | 89 | 28.3 |
| **Medication type** | Metformin | 80 | 25.5 |
|  | Glibenclamide | 32 | 10.2 |
|  | Metformin/Glibenclamide | 202 | 64.3 |

### Dietary knowledge

A total of 146 (46.5%) patients had adequate knowledge regarding the recommended dietary practices for individuals with T2DM, 91 (29.0%) had moderate knowledge, and the remaining 77 (24.5%) had inadequate knowledge.

Three hundred four people (96.8%) were aware that nutrition is crucial in diabetes control. When it came to choosing a proper diet to control blood glucose, 227 (72.3%), 221 (70.4%), 226 (72.0%), 270 (86.0%), and 204 (65.5%) were aware of the importance of eating fruits and vegetables, reducing high fat diet, reducing fried food consumption, minimizing salt consumption, and minimizing sugary foods, respectively. In terms of the immediate influence of diet on blood sugar, 197 (62.7%) recognized that eating little, frequent meals is important, and 206 (65.6%) knew that eating a large piece of food at once may contribute to higher blood sugar. **(Table 3)**

**Table 3:**
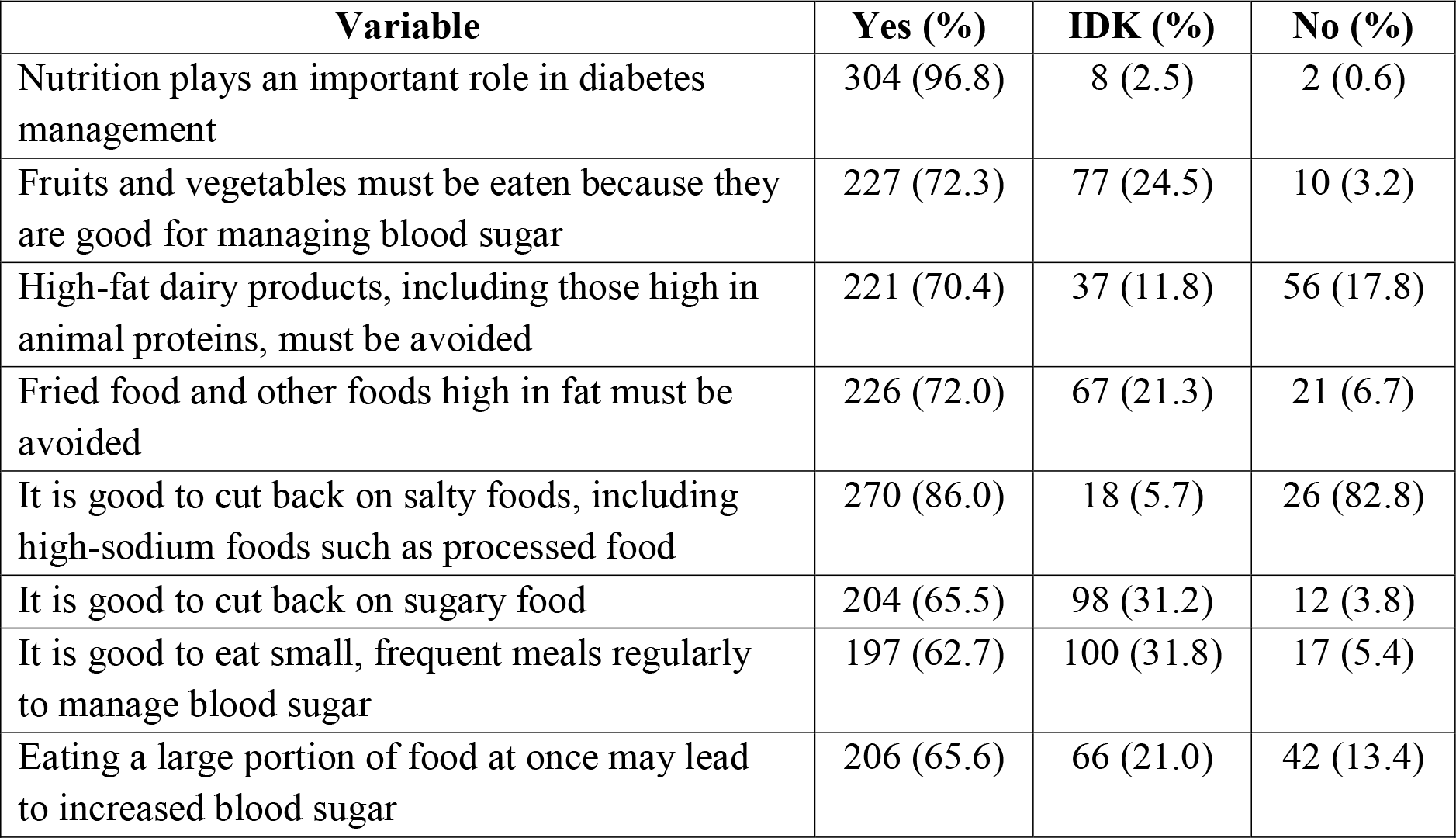
Dietary knowledge of individuals with T2DM on follow up at two tertiary hospitals in Ethiopia, 2023 (n=314)

**Table 4:**
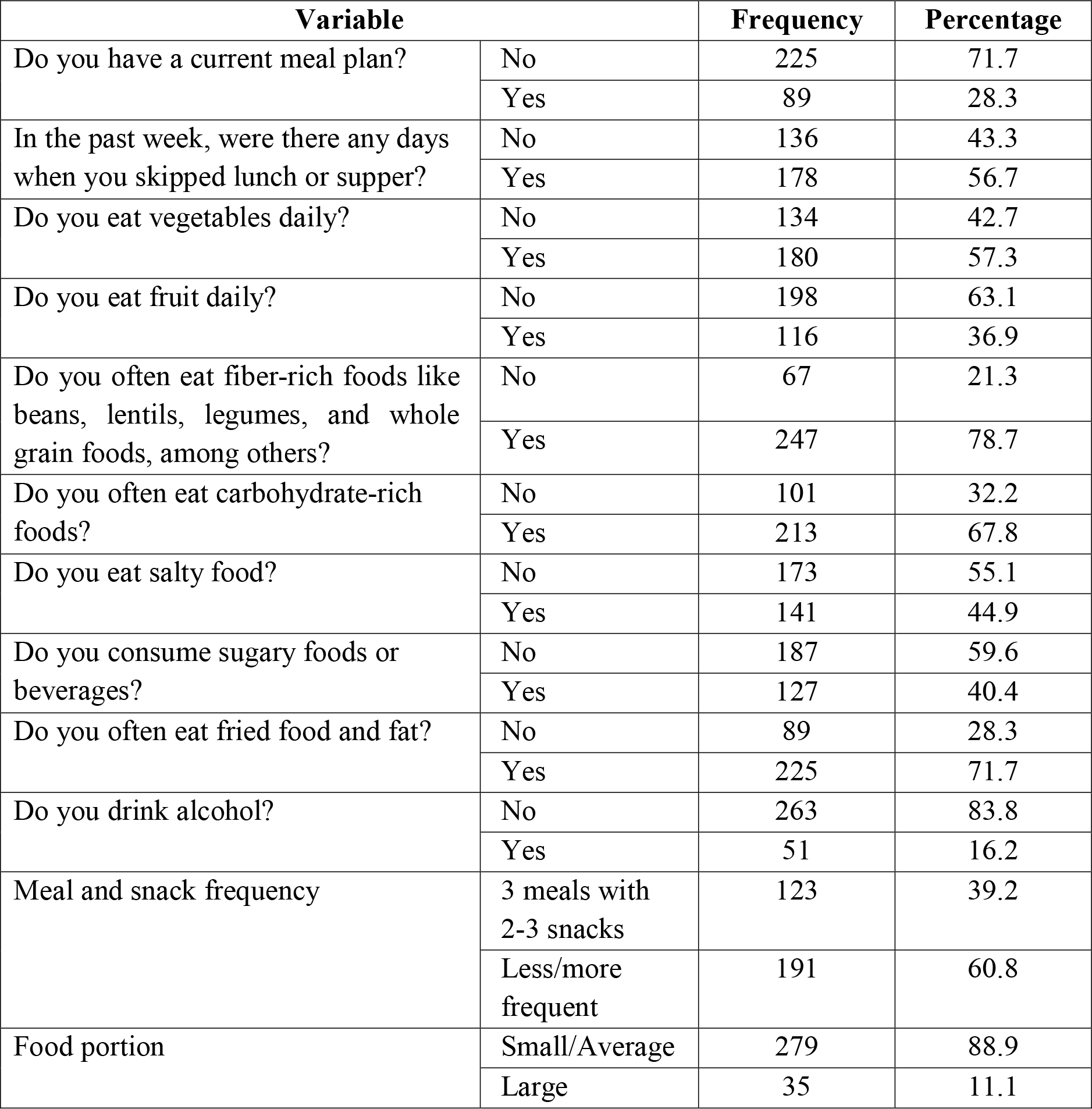
Dietary practice of individuals with T2DM on follow up at two tertiary hospitals in Ethiopia, 2023 (n=314)

| Variable |  | Frequency | Percentage |
| --- | --- | --- | --- |
| Do you have a current meal plan? | No | 225 | 71.7 |
|  | Yes | 89 | 28.3 |
| In the past week, were there any days when you skipped lunch or supper? | No | 136 | 43.3 |
|  | Yes | 178 | 56.7 |
| Do you eat vegetables daily? | No | 134 | 42.7 |
|  | Yes | 180 | 57.3 |
| Do you eat fruit daily? | No | 198 | 63.1 |
|  | Yes | 116 | 36.9 |
| Do you often eat fiber-rich foods like beans, lentils, legumes, and whole grain foods, among others? | No | 67 | 21.3 |
|  | Yes | 247 | 78.7 |
| Do you often eat carbohydrate-rich foods? | No | 101 | 32.2 |
|  | Yes | 213 | 67.8 |
| Do you eat salty food? | No | 173 | 55.1 |
|  | Yes | 141 | 44.9 |
| Do you consume sugary foods or beverages? | No | 187 | 59.6 |
|  | Yes | 127 | 40.4 |
| Do you often eat fried food and fat? | No | 89 | 28.3 |
|  | Yes | 225 | 71.7 |
| Do you drink alcohol? | No | 263 | 83.8 |
|  | Yes | 51 | 16.2 |
| Meal and snack frequency | 3 meals with 2-3 snacks | 123 | 39.2 |
|  | Less/more frequent | 191 | 60.8 |
| Food portion | Small/Average | 279 | 88.9 |
|  | Large | 35 | 11.1 |

**Table 4:** Factors associated with level of glycemic control among individuals with T2DM on follow up at two tertiary hospitals in Ethiopia, 2023 (n=314)

| Variable |  | Glycemic control |  | COR (95% CI) | AOR (95% CI) | p-value |
| --- | --- | --- | --- | --- | --- | --- |
|  |  | Optimal | Sub-optimal |  |  |  |
| **Age category (in years)** | 22-49 | 39 | 35 | 1 | 1 |  |
|  | 50-59 | 44 | 66 | 1.67 (0.92-3.03) | 1.11 (0.49-2.53) | 0.802 |
|  | 60-69 | 16 | 71 | 4.95 (2.43-10.04) | 2.28 (0.87-5.96) | 0.093 |
|  | 70-82 | 8 | 35 | 4.88 (1.99-11.91) | 1.32 (0.36-4.87) | 0.682 |
| **Sex** | Male | 60 | 98 | 1 | 1 |  |
|  | Female | 47 | 109 | 1.42 (0.89-2.27) | 1.02 (0.48-2.19) | 0.952 |
| **Education al level** | Cannot read and write | 7 | 19 | 1 | 1 |  |
|  | Primary | 34 | 109 | 1.18 (0.46-3.05) | 2.38 (0.73-7.71) | 0.149 |
|  | Secondary | 21 | 36 | 0.63 (0.23-1.75) | 2.52 (0.65-9.84) | 0.184 |
|  | Diploma and above | 45 | 43 | 0.35 (0.14-0.92) | 1.96 (0.49-7.89) | 0.342 |
| **Hypertension** | No | 33 | 35 | 1 | 1 |  |
|  | Yes | 74 | 172 | 2.19 (1.27-3.79) | 0.58 (0.26-1.33) | 0.202 |
| **Cardiac disease** | No | 100 | 167 | 1 | 1 |  |
|  | Yes | 7 | 40 | 3.42 (1.48-7.93) | 1.58 (0.57-4.38) | 0.377 |
| **Neurologic conditions** | No | 75 | 113 | 1 | 1 |  |
|  | Yes | 32 | 94 | 1.95 (1.19-3.20) | 0.76 (0.37-1.56) | 0.458 |
| **Renal** | No | 95 | 150 | 1 | 1 |  |
|  | Yes | 12 | 57 | 3.01 (1.53-5.89) | 1.26 (0.51-3.07) | 0.617 |
| **BMI** | <25.0 | 52 | 44 | 1 | 1 |  |
|  | 25.0-29.9 | 42 | 93 | 2.62 (1.52-4.50) | 1.19 (0.55-2.56) | 0.659 |
|  | ≥ 30 | 13 | 70 | **6.36 (3.11-13.01)** | **2.74 (1.05-7.18)** | **0.040\*** |
| **Exercise** | Moderate/vigorous | 46 | 22 | 1 | 1 |  |
|  | Sub-optimal | 61 | 185 | 6.34 (3.53-11.38) | 1.57 (0.62-3.95) | 0.342 |
| **Treatment regimen** | Glibenclamide/Metformin | 77 | 35 | 1 | 1 |  |
|  | Glibenclamide and Metformin | 30 | 172 | **12.61 (7.23-22.01)** | **6.22 (3.05-12.69)** | **<0.001\*** |
| **Dietary practice** | Good | 35 | 169 | 1 | 1 |  |
|  | Moderate | 37 | 31 | 4.19 (1.63-10.74) | 2.65 (0.89-7.84) | 0.079 |
|  | Poor | 35 | 7 | **24.14 (9.92-58.75)** | **7.93 (2.63-23.89)** | **<0.001\*** |

\* Statistically Significant at P<0.05, AOR (Adjusted Odds Ratio), COR (Crude Odds Ratio)

### Dietary practice

Only 42 (13.4%) of the individuals practiced a healthy diet. Sixty-eight (21.7%) had moderate practice and 204 (65.0%) had poor practice.

Eighty-nine (28.3%) had a meal plan and 178 (56.7%) had skipped either lunch or supper within a week of the interview. In terms of daily consumption of nutritious foods, 180 (57.3%) claimed to eat vegetables, 198 (63.1%) eat fruits, and 247 (78.7%) eat fiber rich foods on a daily basis. With regard to avoiding glucose-inducing foods, decreased consumption of carbohydrate-rich foods, salty foods, sugary foods or drinks, and fried foods was practiced by 101 (32.2%), 173 (55.1%), 187 (59.6%), and 89 (28.3%), respectively. The majority (83.3%) do not consume alcohol. One hundred twenty-three (39.2%) had a meal frequency of three meals with two or three snacks, and 279 (88.9%) consumed small to average portions. **(Table 4)**

### Glycemic Control

From the 314 participants, 107 (34.1%, 95% CI= 29.0%-39.2%) had optimal glycemic control and the rest 207 (65.9%, 95% CI= 60.8%-71.0%) had suboptimal glycemic control.

### Factors associated with level of glycemic control

The association of dietary practice and other socio-demographic and clinical characteristics with level of glycemic control was assessed using a binary logistic regression model. From the univariate analysis, age group, gender, educational level, hypertension, heart illness, neurologic disorders, renal disease, BMI, exercise, treatment regimen, and dietary practice were all found to be significant. Then multivariable analysis was then run using these variables.

Accordingly, after controlling for all the confounders in the model, dietary practice was found to be significantly associated with level of glycemic control. The odds of having suboptimal glycemic control among patients with poor dietary practice was 7.93 times higher as compared to those with good dietary practice (**AOR=7.93, 95% CI= 2.63-23.89, p<0.001**). On the other hand, a significant difference in glycemic control level was not observed between those with moderate and good glycemic control.

Furthermore, BMI and treatment regimen were also found to have a significant association with level of glycemic control. Patients with BMI of 30 and above had an increased odds of having suboptimal glycemic control by 2.74 times than patients with BMI of less than 25 (**AOR=2.74, 95% CI= 1.05-7.18%, p=0.04**). Patients taking combination oral anti-diabetic medications (Metformin and Glibenclamide) had 6.22 times increased odds of having suboptimal glycemic control than those taking a single oral anti-diabetic medication of either type (**AOR=6.22, 95% CI= 3.05-12.69, p<0.001**). **(Table 4)**

## DISCUSSION

In this study, we have assessed dietary practice of individuals with T2DM and its association with glycemic control among patients on follow-up at diabetes clinics of two tertiary hospitals in Ethiopia. The study included 314 participants with a median age of 56 years.

The result of this study showed that the majority of individuals with T2DM had either moderate (29.0%) or inadequate (24.5%) knowledge about the recommended dietary practices for their condition. While almost all (96.8%) were aware of the general importance of nutritional intervention in diabetes control, two-thirds were not aware of the specific dietary recommendations. This low level of knowledge could be attributed to inadequate education from their healthcare providers or limited access to additional credible information sources that are prepared in local languages. Furthermore, the dietary practice of the study participants showed that only 42 (13.4%) practiced a healthy diet and 68 (21.7%) had moderate practice. More than three-quarters did not have a meal plan, and more than half did not adhere to the suggested dietary practices. This is also true among individuals who indicated moderate to adequate comprehension of the recommended practice. Despite the fact that most have been living with the disease for an average of 8.6 years, the lower level of knowledge and practice is concerning, especially given that the majority have additional comorbid illnesses that necessitate extra caution in dietary measures as it can result in suboptimal glycemic control, an increased risk of complications, and thus a lower quality of life and increased mortality. Previous studies in similar settings have also shown comparable findings in both knowledge and practice, implying a possible inadequate intervention over time in bringing people to a desirable behavior (15, 25-26).

From the 314 participants, 107 (34.1%) had optimal glycemic control, while the remaining 207 (65.9%) had suboptimal glycemic control. This finding is consistent with previous research conducted in similar setups (8-10). After controlling for potential personal and clinical factors, dietary practice was found to have significant association with glycemic control, with those with poor dietary practice having an almost eight-fold increased odds of having suboptimal glycemic control than those with good dietary practice. This is a little bit of a magnified risk than previous studies conducted in Ethiopia (21,22). This could be due to the characteristics of the study population in our study where majority had additional risks that could increase the risk of suboptimal glycemic control including old age, one or more additional comorbid illnesses (88%), sub-optimal level of physical activity (78%), being overweight (43%) or obese (26%), smoking (4%) and Khat chewing (16%).

Furthermore, BMI and treatment regimen were also found to have a significant association with level of glycemic control, with obese patients and those on combination therapy having almost three times and six times increased odds of having suboptimal glycemic control, respectively. Obesity is known to be associated with increased insulin resistance in the body, making it challenging to achieve treatment goals even when the patient is on appropriate medication. Additionally, being on combination therapy indicates that the threshold for controlling blood glucose is high, which could be due to different personal and clinical factors interfering with optimal control, as well as the possibility of poor drug adherence with increased pill burden, making glycemic control difficult in these individuals. In previous studies as well, including systematic reviews and meta-analyses, these characteristics have been demonstrated to be associated with suboptimal glycemic control (10, 27-29).

The study was carried out at the country’s two main tertiary hospitals, which have a high patient flow and are supposed to deliver the finest available comprehensive management. As a result, the findings of this study enable us to better understand the gap among patients under this type of care and plan accordingly. However, the study did not include additional characteristics such as treatment adherence and other concomitant illness control levels which might contribute to the level of glycemic control in addition to the dietary practice and other factors studied.

## CONCLUSIONS

The study revealed that dietary knowledge and practice among individuals with T2DM are very low, as is the level of suboptimal glycemic control, which are similar to studies conducted years back, indicating a lack of improvement in the desired behavior over time. Poor dietary practice was significantly associated with suboptimal glycemic control. The findings of this study highlight the importance of targeting interventions that enhance the understanding and application of dietary practice in individuals with T2DM, including strengthening patient education using culturally sensitive tools and strategies. Furthermore, clinicians should be cautious in the management of patients with the identified significant factors that lead to suboptimal glycemic control.

## Data Availability

All relevant data are available upon reasonable request.

## DECLARATION

### Ethical Considerations

The study was conducted after obtaining ethical clearance from the Ethics Review Committee of St. Paul’s Hospital Millennium Medical College and Addis Ababa University College of Health Sciences. Eligible participants patients were given clear information about the purpose of the study and written informed consent was obtained from all. To ensure confidentiality, name and other identifiers of patients were not recorded on the data collection tools. Access to the collected information was limited to the research team and confidentiality was maintained throughout the project.

### Availability of data and materials

All relevant data are available upon reasonable request.

### Competing interests

The authors declare that they have no known competing interests

### Funding source

This research did not receive any specific grant from funding agencies in the public, commercial, or not-for-profit sectors.

### Author’s Contribution

FAK conceived and designed the study. TWL, MZM, IAY, BHT, YSS, BST, MFB, and DSW contributed to the conception and design of the study. FAK and TWL performed statistical analysis, and drafted the initial manuscript. MZM, IAY, and BHT contributed to the statistical analysis and interpretation of the findings. YSS, BST, MFB, and DSW revised the manuscript. All authors approved the final version of the manuscript.

## Acknowledgement

The authors would like to thank the data collectors for their hard work and the hospitals for facilitating the research work.

## Notes

### Competing Interest Statement

The authors have declared no competing interest.

